# Ultraviolet A Radiation and COVID-19 Deaths in the USA with replication studies in England and Italy

**DOI:** 10.1101/2020.07.03.20145912

**Authors:** Mark Cherrie, Tom Clemens, Claudio Colandrea, Zhiqiang Feng, David J Webb, Chris Dibben, Richard B Weller

## Abstract

Seasonal variation in environmental meteorological conditions affect the incidence of infectious diseases. Ultraviolet A (UVA) radiation induces release of cutaneous photolabile nitric oxide (NO) impacting the cardiovascular system and metabolic syndrome, COVID-19 risk factors. NO also inhibits the replication of SARS-CoV. We therefore model the relationship between UVA radiation, derived from remote sensed data, and COVID-19 deaths for counties across the USA during their ‘UV vitamin D winter’ (Jan-April) adjusting for confounding including by temperature and humidity. The Mortality Risk Ratio (MRR) falls by 29% (40% -15% (95% CI)) per 100 (KJ/m^2^) increase in mean daily UVA. We replicate this in independent studies in Italy and England and estimate a pooled decline in MRR of 32% (48%-12%) per 100 KJ/m^2^ across the three studies.

## Introduction

Seasonality(1) and variation in temperature(2), humidity(3) and UV radiation(4) are related to the incidence of several infectious diseases. COVID-19 arose less than 6 months ago, and it is thus not possible to describe seasonal variation. Nonetheless, spatial variation in levels of environmental UV in the early pandemic allows its relationship with COVID-19 mortality to be modelled. We have previously described a novel NO driven, vitamin D independent, mechanism(5), by which sunlight can lower blood pressure, and at the population level we have shown that UV is associated with lower blood pressure(6) and a reduced incidence of myocardial infarctions(7). The same UV driven mechanism may also cause seasonal variation in development of diabetes and metabolic syndrome(8). Given the apparent greater severity of illness and risk of death amongst those with these conditions(9, 10) and the importance of season, we investigate if ambient UVA exposure is associated with COVID-19 deaths across the USA and is the finding replicated in studies of England and Italy.

## Results

Daily mean UVA (January-April 2020) varied between 450-1,000 KJ/m^2^ across the three countries, with lower average levels experienced across England (Figure 1 a,b,c). Our fully adjusted model shows reductions in Mortality Risk Ratios (MRR) of 0.71 in the USA per 100 increase in UVA (KJ/m^2^) (Table 1). We found a similar size of effect in our two replication studies; an MRR in Italy of 0.81 and in England 0.49 with a pooled estimate of 0.68 (Figure 2d). This represents a halving of the average risk of death across the lower and narrower range of UVA experienced across England and across the higher and wider range across Italy and the USA (Figure 2 a,b,c).

**Table 1:**
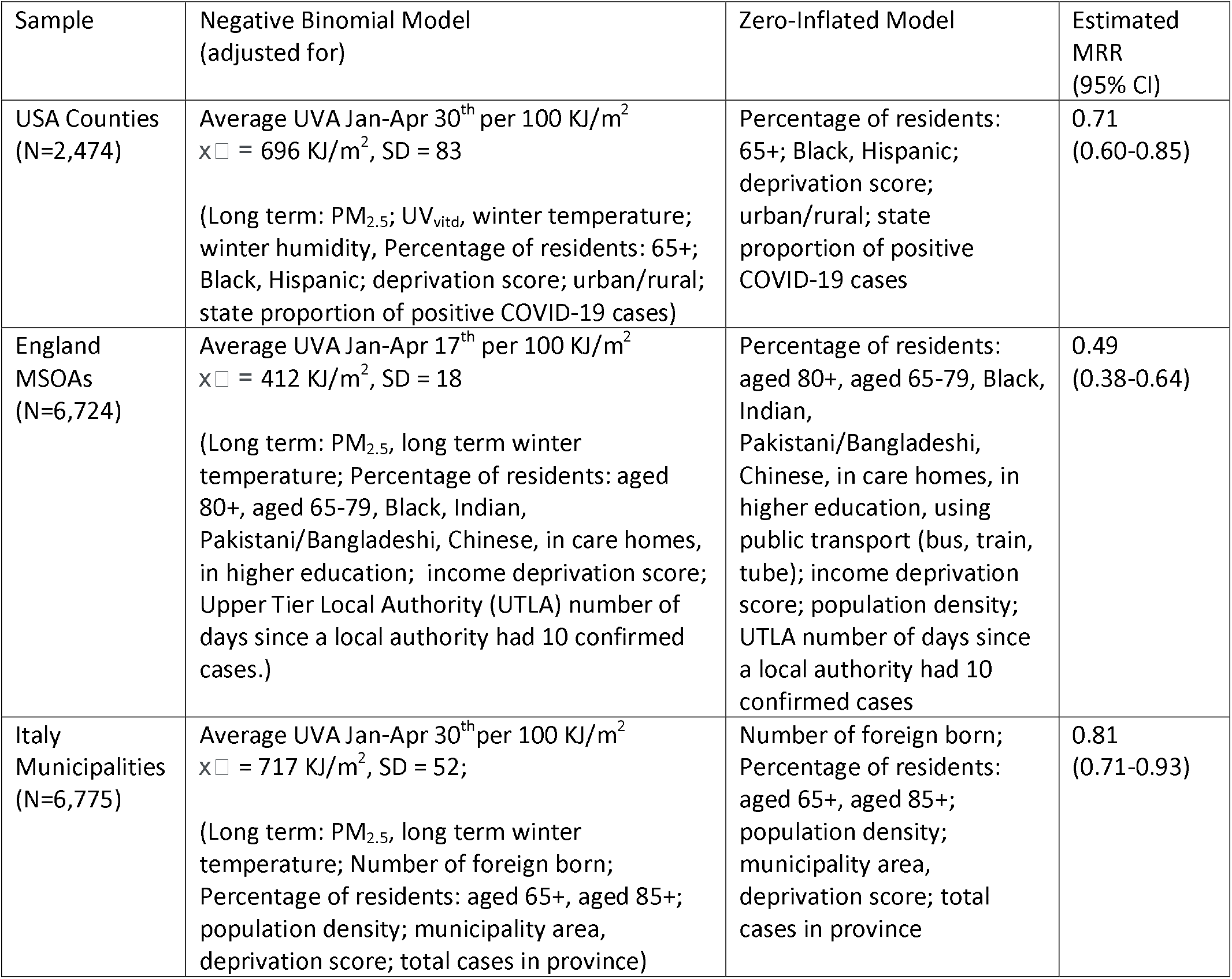
Model estimates for the relationship between UVA and COVID-19 death

**Figure 1:**
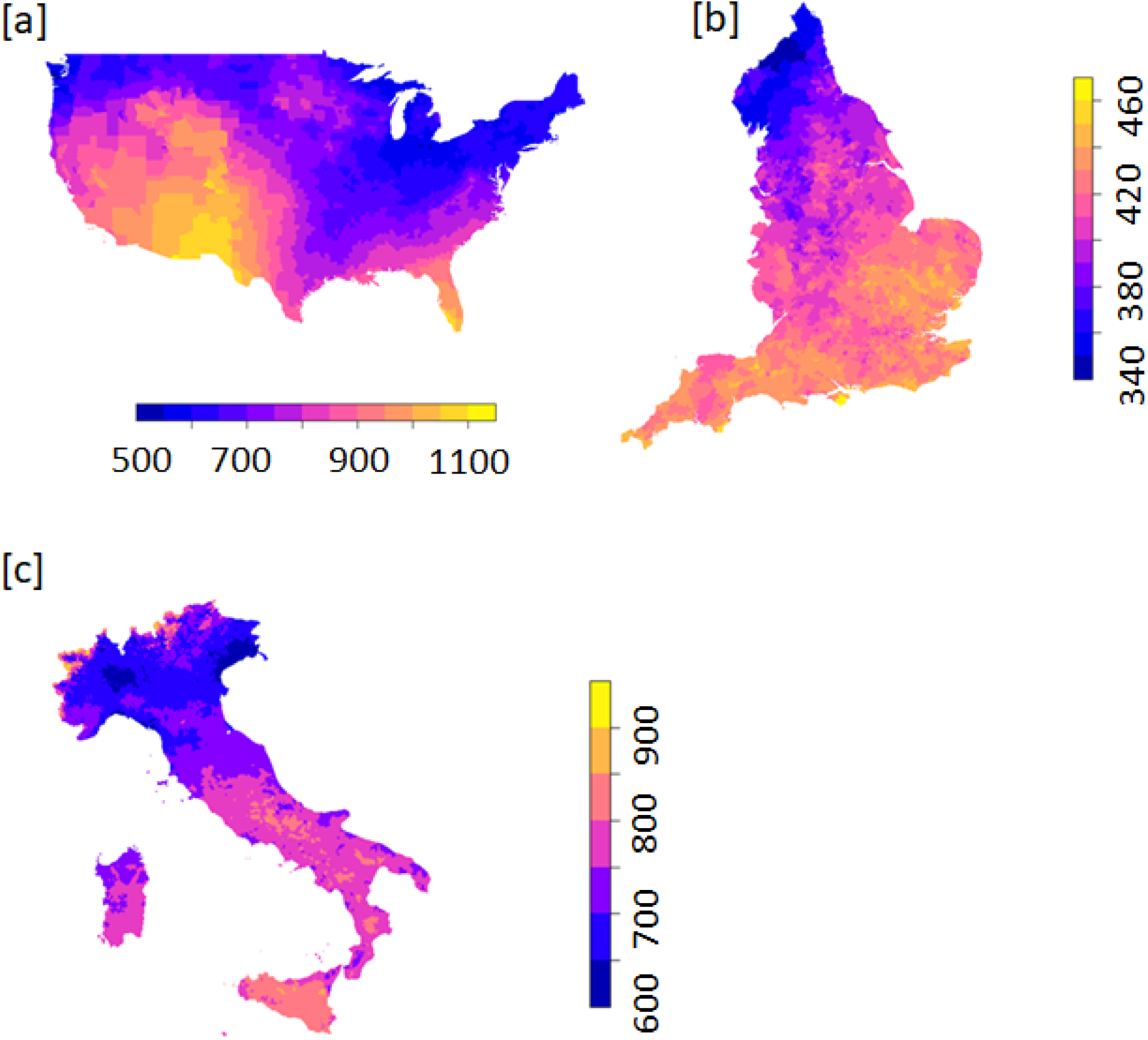
Average daily mean UVA (KJ/m^2^) Jan-April [a] USA [b] England [c] Italy

**Figure 2.**
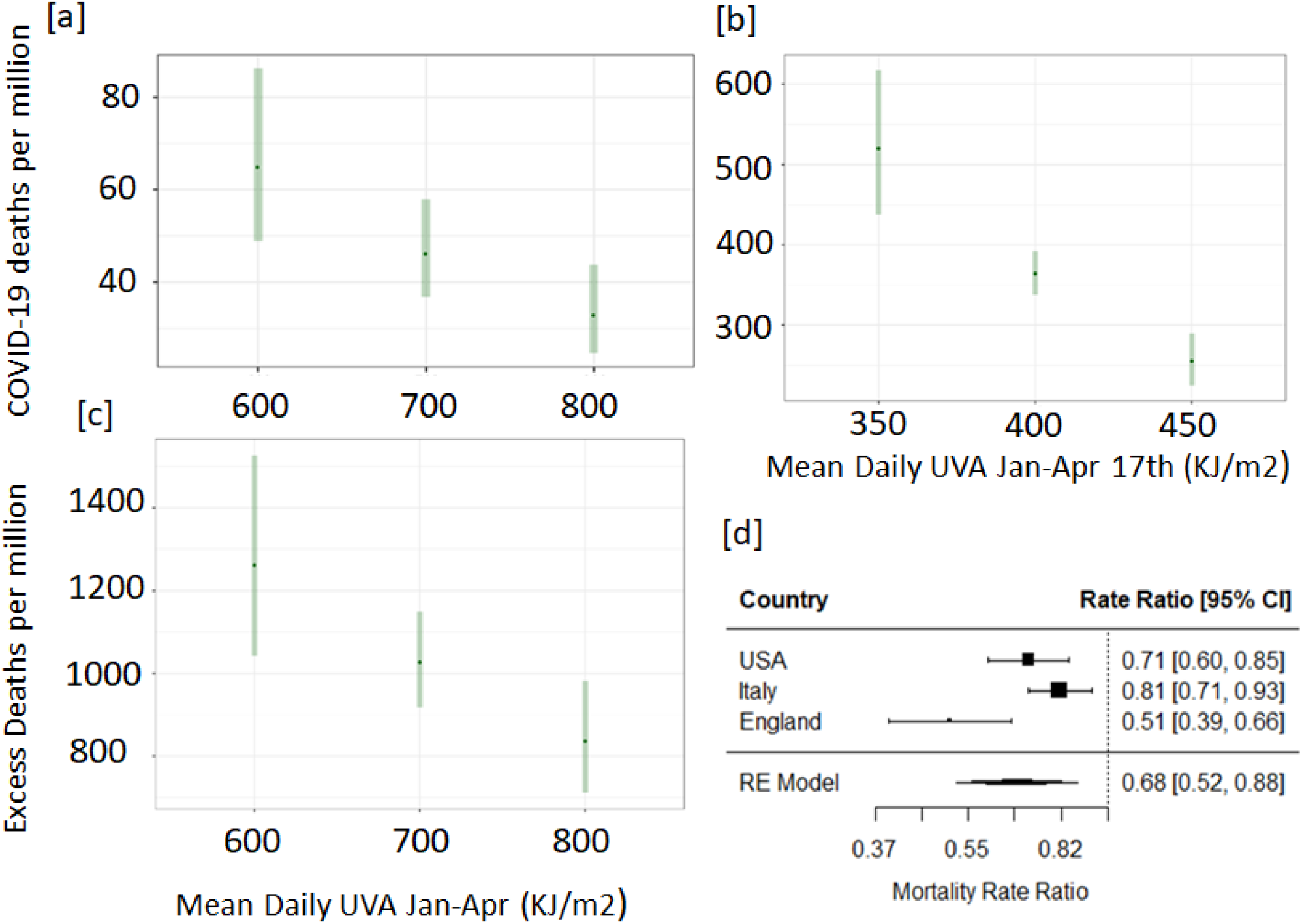
Predicted COVID-19 rates of deaths at selected levels of UVA in the [a] USA [b] England [c] Italy, given the model random effect, at the mean level of all other covariates. [d] MRR per 100 KJ/m^2^ increase in mean daily UVA – pooled estimate from random effects model.

## Discussion

Our analysis suggests that higher ambient UVA exposure is associated with lower COVID-19 specific mortality. This effect is independent of temperature, humidity and UV within the vitamin D action spectrum.

Seasonal variations in disease incidence can be caused by environmental, behavioural, and immunological factors with the relative importance of these varying by location and disease. UV may have a direct effect on the viability of SARS-COV-2 virus in airborne droplets and on fomites, thus reducing both infection rates, and also the size of inoculum in those becoming infected, with correspondingly reduced disease severity (11, 12). UVA radiation photo-releases nitric oxide (NO) from stores in the skin whence it is mobilised to the systemic circulation, causing vasodilation and reduction in blood pressure(5), offering cardiovascular and metabolic benefits from UV exposure(5, 8). As cardio-metabolic disease and possibly hypertension(13) increase the risk of death from COVID-19, any UV driven improvements in these risk factors would be expected to reduce mortality(14). Nitric oxide may also have a specific effect on COVID-19. It inhibits replication of SARS CoV(15), probably by S-nitrosating the spike protein, thus preventing the post translational palmitoylation of the spike protein, required for fusion with its cognate angiotensin converting enzyme 2 receptor (ACE2R) (16). The spike protein of SARS CoV is highly homologous to that of SARS CoV2(17, 18) suggesting that NO may similarly limit binding to ACE2R by SARS CoV2 with reduction in disease transmission and severity. Endothelial damage, with reduced homeostatic endothelial NO synthase activity may underlie widespread organ involvement(19). This would be mitigated by photochemical NO production. Tolerance to UVA could be explained by increasing melanin in the skin, which blocks UV penetration; approximately half of the annual variation in forehead pigmentation for Caucasian individuals living in Copenhagen (55.68°N) occurred between January and April (20).

We believe we have adjusted our models for the clinically significant factors that might be spatially and temporally associated with UVA in our studies but if any others exists, they might plausibly explain the relationships identified. UVA and covariates are measured at the small area level, there could be misclassification of deaths and rates of infection are estimated within the model and with indirect measures. However, any resulting measurement errors seem unlikely to be correlated with spatial variation in UVA and therefore biasing. The random effect in our models will incorporate differences between socially and politically distinct regions (States, Local Authorities and Municipalities) that might induce a spurious relationship between UVA and mortality. The replication of the findings across three countries with very different health systems, economic and political structures, pandemic situations and climates suggest a robust finding.

This study is observational and therefore any causal interpretation needs to be taken with caution. However, if the relationship identified proves to be causal, it suggests that optimising sun exposure may be a possible public health intervention. Given that the effect appears independent of a vitamin D pathway, it suggests possible new COVID-19 therapies and the importance of exploring the role of circulating NO.

## Materials and methods

We model COVID-19 classified deaths in the USA across 2,474 Counties, for January-April 2020. We only include counties that were experiencing levels of UV too low (equivalent to a monthly mean UV_vitd_ of under 165 KJ/m^2^) to be inducing significant levels of cutaneous vitamin D3 synthesis during the study period (‘UV vitamin D winter’). We derive UVA measures from remote sensing data for the same period for each county and estimate, in a multilevel zero-inflated negative binomial (ZINB) model, their relationship with COVID-19 mortality with a random effect for States. The ‘at-risk’ population is the total county population, with [1] the higher level random effect, [2] a measure of the proportion of population tested positive for COVID-19 at the state level and [3] measures of infection susceptibility (in both the ZI and NB parts of model), used to incorporate viral spread. We then attempted to replicate this model for excess deaths (deaths over 2015-19 period average) in 6,775 Municipalities in Italy and for COVID-19 related deaths in 6,724 small areas across England. The models in each country are adjusted for potentially confounding factors at the small area level (table 1) and were independently specified by sub-groups in the research team. All analyses were rerun using a negative binomial formulation and no difference in broad findings were identified. We use meta-analysis to produce a pooled overall estimate using a random effects model.

The UVA dataset was produced by JAXA (Japan Aerospace Exploration Agency) using the MODerate resolution Imaging Spectroradiometer (MODIS) instrument. Atmospheric absorption due to the ozone and water vapour were accounted for. Downward irradiance values (i.e. combined direct and diffuse radiation on a horizontal plane) for UVA (315nm-400nm) were converted to daily values by using the diurnal cycle of solar zenith angle with instantaneous atmospheric conditions.

All data software code, and other detailed methods available here: https://github.com/markocherrie/COVID19_UVA

## Data Availability

Data Availability
The UVA estimates were developed by JAXA, with MODIS data provided by NASA GSFC, and available from ftp://apollo.eorc.jaxa.jp/pub/JASMES/Global_05km/. The long term UVvitd estimates were developed by NCAR and available from https://www2.acom.ucar.edu/modeling/tuv-download. The USA COVID-19 death and county level indicators of population, socioeconomic and environmental data (PM2.5 and temperature) were developed previously by Xiao Wu et al., and are available here: https://github.com/wxwx1993/PM_COVID.
Code Availability
The code and data to reproduce the current study is held at https://github.com/markocherrie/COVID19_UVA.

ftp://apollo.eorc.jaxa.jp/pub/JASMES/Global_05km/

https://www2.acom.ucar.edu/modeling/tuv-download

https://github.com/wxwx1993/PM_COVID.

## Acknowledgements

MC was supported by Health Data Research UK (HDR-5012).

CC was supported by the European Union’s Horizon 2020 research and innovation programme Grant agreement No. <u>676060</u>

## Extended methods

### Study Setting and Participants

We used an ecological regression approach to model COVID-19 deaths in small areas (counties) across the contiguous⍰SA during the early part of the COVID-19 pandemic (January to April 2020). Our main analysis was for USA counties (N=2,474). We then carried out replication studies for COVID-19 deaths across English Middle Layer Super Output Areas (MSOAs) (N=6,724) and excess deaths across Italian municipalities (N=6,775). We only included ‘small areas’ that were experiencing levels of UV too low to be inducing significant cutaneous vitamin D3 synthesis at any time during the study period (‘UV vitamin D winter’), to reduce potential confounding through a UVB vitamin D pathway. Too low was defined as a monthly mean UV on the 252-330nm spectrum (the Vitamin D active spectrum - **UVvitd**) of under 165 KJ/m2 ^i^. This meant that 2474 counties (out of 3088) in the USA were within the analysis (Figure S1).

All analyses were undertaken in R 3.6.1.

**Figure S1:**
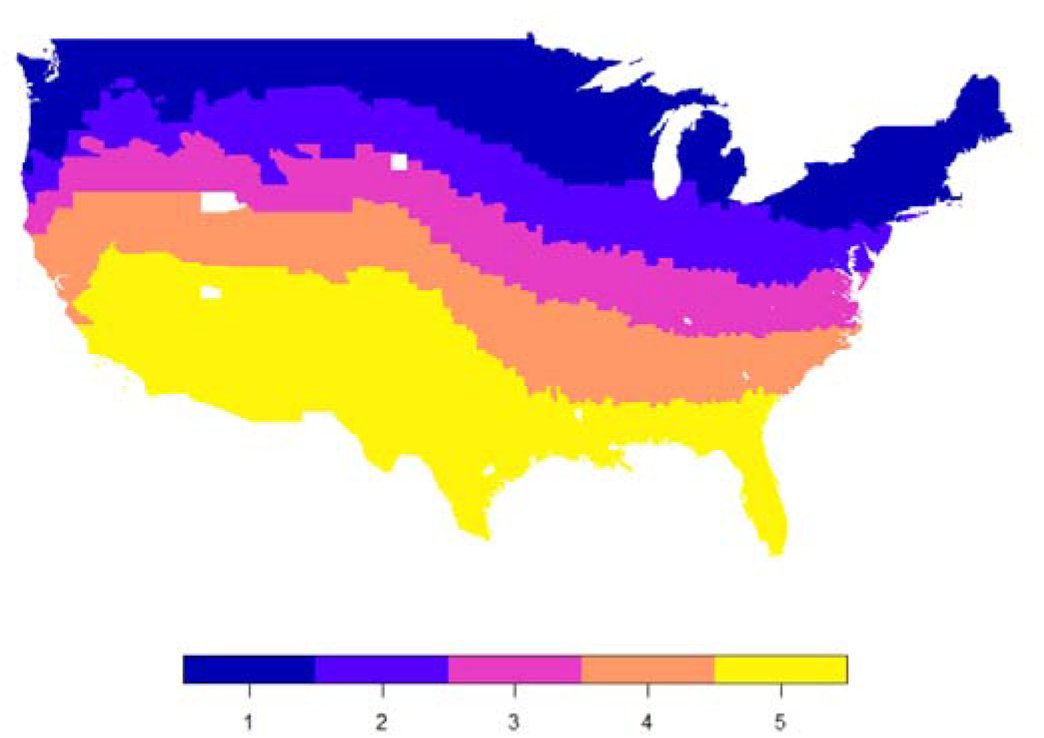
Counties that were excluded from the study because they had monthly mean UVvitd of over 165 KJ/m2 are shown in yellow.

### Outcome measure

USA COVID-19 deaths were drawn from data compiled by the Center for Systems Science and Engineering at Johns Hopkins University. We included deaths occurring between January 22nd and April 30^th^ 2020. These data were derived from death certificates, with information on cause and circumstances of death recorded, collected by the Center for Disease Control and Prevention (CDC). A COVID-19 death represents a case where the practitioner suspected that COVID-19 played a role in the death, even if it was not directly attributable to the death. English COVID-19 deaths were drawn from data compiled by the UK Office for National Statistics^ii^. Data were extracted for March 1^st^ to April 17^th^ 2020. Deaths were included in this dataset if COVID-19 was mentioned on the death certificate, with a delay of usually five days between occurrence and registration. In Italy there is no COVID-19 classified mortality data available for municipalities. Instead, we had to estimate this from excess deaths. Italian excess deaths are drawn from ISTAT (Italian Institute of Statistics)^iii^. These data are only available for 92% of municipalities (7,270/7,904). Data were for the period of March 1^st^ to April 30^th^ for 2015-2019 and 2020. We classified excess deaths as the positive difference in deaths between 2020 and 2015-19, for the same period, with negative values recoded to zero.

### Ambient UV data

We derived mean daily UVA for the small-areas in each study – USA (Jan 1^st^ – April 30^th^); England (Jan 1^st^ – April 17^th^) and Italy (Jan1^st^ – April 30^th^). We start our observation of UVA before the period in which we were recording deaths because we believed the protective effect of UVA, if it existed, might occur sometime before an observed deaths possibly during the initial infection period or even before this. The UVA dataset was produced by JAXA (Japan Aerospace Exploration Agency) using the MODerate resolution Imaging Spectroradiometer (MODIS) instrument on board NASA’s Aqua and Terra satellites^iv^. Atmospheric absorption due to the ozone and water vapour (cloudiness) were accounted for by using a simplified planetary atmosphere (clear atmosphere positioned above a cloud layer). Downward irradiance values (i.e. combined direct and diffuse radiation on a horizontal plane) for UVA (315nm-400nm) were converted to daily values by using the diurnal cycle of solar zenith angle with instantaneous atmospheric conditions. These data are available to download at a 5km by 5km spatial resolution. UVA data were aggregated for USA counties, English MSOAs and Italian municipalities and expressed as mean daily KJ/m^2^ . The R ‘velox’ package was used to extract raster values for each polygon. The function extracts all the UVA values from the UV raster cell centroids that intersect with the county polygon. The mean of the extracted UVA values was then assigned to each county polygon. This procedure is used for most of the small areas (given that the county boundary included at least one UV raster cell centroid). However, in a few cases, when the county polygon was small (or oddly shaped) and did not intersect with any cell centroid, we extracted values based on the small county polygon intersecting with the entire UV raster cell.

A long term UV_vitd_ dataset (30-year monthly average) developed by the National Center for Atmospheric Research (NCAR) using the Tropospheric Ultraviolet and visible (TUV) radiation model was used^v^. This model uses the Total Ozone Mapping Spectrometer (TOMS) on board several satellites (Nimbus-7, Meteor-3 and Earth Probe) to account for atmospheric ozone and climatological cloudiness (defined by TOMS reflectivity at 380 nm). Mean monthly values at a 1° (latitude) by 1.25° (longitude) spatial resolution are available for the period 1979-2000. UV_vitd_ data were aggregated for US counties and expressed in mean monthly KJ/m^2^. We used the highest quintile as the cut off for year-round vitamin D synthesis, which corresponds to a monthly mean of over 165 KJ/m^2^. It also corresponds approximately to the 37°N parallel; Holick suggests that people living North of this latitude will not receive sufficient UV for vitamin D synthesis in the winter^3^.

### Covariates

A number of demographic, socioeconomic, long term environment exposures and variables to measure infection susceptibility were measured and used in our models. This was to appropriately adjust for spatial associations, with both UVA and COVID-19 mortality, which might otherwise lead to a spurious relationship between UVA and COVID-19 mortality. These are all measured at the small area level. The selection of the covariates was made independently by different members of the research team from different sets of available data, to maintain a level of independence between the main study and the replications, but with the same goal of appropriate adjustment.

### Demographic

Older age and ethnicity were associated with higher risk of COVID-19 death, possibly due to higher prevalence of comorbidities, including hypertension, heart disease and respiratory diseases^vi^. We used data on county percentage of older residents (≥ age 65), percentage of Hispanic residents and percentage of Black residents for the USA to measure risk factors associated with age and ethnicity in the USA. We used data from ONS mid-year population estimates of 2018 on percentage of residents who are: aged 80 or over, aged 65-79, the 2011 census on living in care homes, Black, Indian, Pakistani/Bangladeshi and Chinese in England. We used data from ISTAT on number of foreign-born residents and the percentage of residents: aged 65 and above and aged 85 and above, for Italy.

### Socioeconomic Deprivation

Poorer citizens are at higher risk of infection due to essential working and death due to pre-existing health conditions^vii^. Socioeconomic deprivation is defined in the USA by the first principal component score from a Principle Component Analysis of the following county variables: percentage in poverty, median house value, median house income, percentage owner occupied and percent of population with less than a high school education. We reversed the direction of percentage in poverty and percent of population with less than a high school education so that higher score represented higher affluence. To capture socioeconomic deprivation in England we used percentage of residents under 21 who did not enter higher education and an income deprivation score indicating the percentage of people who received low income benefits^viii^. For our Italian study, we used the Italian Deprivation Index, calculated by ISFOL (an Italian research institute), which considers income, education, living conditions, unemployment and household composition.

### Long term environment

Higher PM_2.5_ is linked with a range of respiratory and cardiovascular disease and shown to increase COVID mortality rate in other analyses^ix^. Long term PM_2.5_ (2000-2016) data at a 0.01° by 0.01° resolution were modelled using satellite and monitored PM_2.5_ station data^x^. We used these data for both the USA and Italy. In England, long term 2014-2018 PM_2.5_ at a 1km by 1km resolution was modelled using monitored PM_2.5_ station data^xi^. Variation in temperature is associated with COVID-19 mortality^xii^. Long term mean monthly winter temperature (Dec-Feb) at a 4km by 4km resolution for 2000-2016, was modelled using satellite data^xiii^ for the USA. Long term mean monthly winter temperature (Dec-Feb) at a 1km by 1km resolution for 1981-2010 was modelled using interpolation of Meteorology Office weather stations for England^xiv^. Long term median land surface temperature (Dec-Feb) daytime monthly median value at a 1km by 1km resolution for 2000-2017, was modelled using satellite data for Italy^xv^.

### Viral exposure

Exposure to SARS-CoV-2 virus is the principle risk factor for a COVID-19 related death. The population ‘at-risk’ therefore needs to be adjusted for exposure in case difference in factors increasing or decreasing risk of exposure are associated with spatial variance in UVA levels. In densely populated, urban or peri-urban areas, with high use of public transport COVID-19 transmission is faster and the prevalence of cases higher. Probable exposure is therefore estimated through county population density, urban/rural status and state percentage of positive COVID-19 tests in the USA. We used population density from the 2018 mid-year population estimates of ONS, percentage of residents using different forms of transport (bus, train, tube) from the 2011 census and Upper Tier Local Authority (UTLA) number of days since a local authority had 10 confirmed cases in England. We used ISTAT population density from 2019, the municipality area, and total cases in province in Italy up to the 30^th^ of April^xvi^. Our adjustment for measures or estimates of cases of COVID-19 at the higher geographical level (i.e. State, UTLA, and Province) may represent a situation of ‘over control’ because it is possible that UVA will reduce the levels of the virus in environmental circulation. However, one would still expect the model to appropriately measure a true effect for the small areas within the higher-level geography after this adjustment.

## Statistical Analysis

### Overview

The dependent variable in our analysis is counts of deaths due to COVID-19 in small geographical areas in the USA and England and counts of excess death in Italy. Because the counts of deaths will be driven, in the early stages of a contagious disease pandemic, by a risk that is likely to vary spatially, the mean counts of deaths across small areas are likely to be much smaller than the variance of the counts between the small areas (i.e. where there are outbreaks and high transmission in a few places and many places with no or very little exposure to the virus) and there will be more zeroes than you would expect given a Poisson process. This means that the assumptions for a Poisson model, the usual approach for counts of death, will not be met. In this analysis we therefore use a zero inflated negative binomial (ZINB) model. This not only handles appropriately the mean and variance not being equal but also the preponderance of zero counts, in this instance due to the fact that a large number of areas will have had no or little exposure to the virus and therefore no risk of death due to it. The ZINB approach models zero counts of death as two different processes: [1] no exposure to the virus, [2] protection from death when exposed. The zero inflated part (ZI) models the likelihood of exposure (a logit model) and the negative binomial (NB) part models the hypothesised protective effect of UVA exposure and adjustment for potential confounders. The variables we used in the zero inflated part are proposed risk factors for exposure to the virus and are listed in table 1 in the manuscript and are discussed above in this supplement.

We included a random effect in the model. This was a random intercept for a higher and administratively important geographical unit in each country. In the USA this was the State, in England the Local Authority and in Italy the Province. This random effect had two purposes. First, it captures the systematic way the risk of death from COVID19 might be related to this higher geography. This might for example be due to differences in political administration, health services, funding, public health effects or levels of infection. Without adjusting for this, any association between the higher-level geography and UVA might be confounding. Second, this random effect meant that the standard errors associated with the model estimates are robustly calculated, taking into account spatial clustering within the datasets associated with the higher geographical level.

As a sensitivity test, all models were re-estimated using an NB model with a random effect as above (i.e. without a zero-inflation part) and a comparison made with the two sets of results. Each of the country models were specified independently by separate team members.

All models were fitted using the glmmTMB package for R^xvii^ which fits random effect generalised linear models described in Bolker *et. al*.^xviii^

Main Model (USA)

We therefore estimated for the USA, in a multilevel ZINB model (using the glmmTMB package), the relationship between ambient UVA and COVID-19 deaths (21^st^ January-April 30^th^), with a state level (N=46) random effect. The ‘at-risk’ population was the total county population, with the [1] state level random effect, [2] a measure of the proportion of population tested positive for COVID-19 at the state level and [3] measures of infection susceptibility (county population density, urban-rural status), used to incorporate viral exposure into the model (in effect ‘correcting’ the at-risk population to be the exposed population not the entire population). The NB model was adjusted at the county level for: percentage of older residents (≥ age 65), Hispanic and Black residents; socioeconomic deprivation and long term modelled 2000-2016 PM_2.5_, long term mean winter temperature (Dec-Feb) and long term mean winter humidity (Dec-Feb) to remove any potential confounding by spatially associated risk factors. The ZI model included: Percentage of residents: 65+; Black, Hispanic; deprivation score; urban/rural; state proportion of positive COVID-19 cases. The ZI part of the model incorporated: percentage of residents: 65+; Black, Hispanic; deprivation score; urban/rural; state proportion of positive COVID-19 cases.

### Replication Model 1 (England)

Our first replication of the USA model was a model of the relationship between ambient UVA and COVID-19 deaths in England (1^st^ of March to 17^th^ of April), with random effects for Upper Tier Local Authorities (UTLA) (the main level of local government in the UK) (N=150). The ‘at risk’ population was the total Middle Super output Area (MSOA – a statistical area unit of population average size 7,200) population, with the [1] UTLA level random effect, [2] number of days since a UTLA had 10 confirmed cases, and [3] measures of infection susceptibility (MSOA population density, population using public transport – bus, train and tube). The NB model was adjusted at the MSOA level for: Long term PM2.5, long term winter temperature, percentage of residents: aged 80+, aged 65-79, Black, Indian, Pakistani/Bangladeshi, Chinese, in care homes, in higher education and income deprivation score. The ZI model included: Percentage of residents: aged 80+, aged 65-79, Black, Indian, Pakistani/Bangladeshi, Chinese, in care homes, in higher education, using public transport (bus, train, tube); income deprivation score; population density; UTLA number of days since a local authority had 10 confirmed cases.

### Replication Model 2 (Italy)

Our second replication of the USA model was a model of the relationship between ambient UVA and excess deaths in Italy (1^st^ of March to 30^th^ of April), with Province (N=104) level random effects. The ‘at risk’ population was the total municipality population, with the [1] province level random effect, [2] number of tests for COVID-19 cases at the Province level and [3] measures of infection susceptibility (municipality population density and area). The NB model was adjusted at the municipality level for: Long term PM_2.5_, long term winter temperature, Number of foreign-born, Percentage of residents: aged 65+, aged 85+ and deprivation. The ZI model included: Number of foreign-born; Percentage of residents: aged 65+, aged85+; population density; municipality area, deprivation score; total cases in province.

### Meta-analysis

Finally, we carried out a meta-analysis to estimate the pooled effect across the three studies. Using a random-effects model, the UVA coefficient from the three ZINB model (with random effects) studies were assumed to be a random sample from a hypothetical large collection of all such possible studies. Because we were interested in estimating the ‘true’ effect of UVA on COVID-19 survival, as it might occur anywhere, this was felt to be the appropriate specification. In the UK and USA, we counted a reasonably ‘direct’ measure of COVID-19 deaths. In Italy, because of an absence COVID-19 specific mortality figures, we had to use excess deaths. This would naturally include other deaths that may have been due to the effect of COVID-19 but not directly attributable to the virus. For example, deaths that may have occurred due to say septic shock in patients who would have been admitted to ITU (and may have survived under ‘normal conditions’) but didn’t because the system was overwhelmed. However, we do not feel this would have biased the estimate of the UVA estimate because these ‘extra’ non-COVID-19 seemed unlikely to be strongly associated with spatial variation in UVA exposure and would be mostly related to the dominating impact of COVID-19 prevalence in the community which was likely to driven by other factors. We used a restricted maximum-likelihood estimator with no adjustments.

### Software

Effect estimates are presented as mortality rate ratios with 95% confidence intervals. We predicted the number of deaths per million population at suitable levels of UVA by calculating the marginal means in the ‘emmeans’ package in R^xix^. To calculate the average ‘true’ effect of UVA on COVID-19 related death in all similar countries to those included, we used a random effects model as part of the ‘metafor’ package in R^xx^ to calculate a cross-county pooled estimate of the MRR.

## Data and code availability

The data and code to reproduce the current study can be found here: https://github.com/markocherrie/COVID19_UVA.

## Notes

### Competing Interest Statement

The authors have declared no competing interest.

### Funding Statement

No external funding

### Author Declarations

This study used publicly available datasets and there was no link to individual patients

### Summary of Updates

We have submitted the text, including supplementary material which has been under review at PNAS since July 2020 (today's date 22nd December).

